# Epidemiology and transmission of COVID-19 in cases and close contacts in Georgia in the first four months of the epidemic

**DOI:** 10.1101/2021.03.22.21254082

**Authors:** Josephine G. Walker, Irine Tskhomelidze, Adam Trickey, Vladimer Getia, Lia Gvinjilia, Paata Imnadze, Tinatin Kuchuloria, Aaron G. Lim, Jack Stone, Sophia Surguladze, Maia Tsereteli, Khatuna Zakhashvili, Peter Vickerman, Amiran Gamkrelidze

**Affiliations:** Population Health Sciences, University of Bristol, Bristol, UK; The Task Force for Global Health,Tbilisi, Georgia; National Center for Disease Control and Public Health, Tbilisi, Georgia; NIHR Health Protection Research Unit in Behavioural Science and Evaluation, University of Bristol, Bristol, UK

## Abstract

**Background:** Between February and June 2020, 917 COVID-19 cases and 14 COVID-19-related deaths were reported in Georgia. Early on, Georgia implemented non-pharmaceutical interventions (NPI) including extensive contact tracing and restrictions on movement.

**Aim:** To characterize the demographics of those tested and infected with COVID-19 in Georgia; to evaluate factors associated with transmission between cases and their contacts; and to determine how transmission varied due to NPI up to 24 June 2020.

**Methods:** We use data gathered by the Georgian National Center for Disease Control on all polymerase chain reaction tests conducted (among symptomatic patients, through routine testing and contact tracing); hospitalization data for confirmed cases, and contact tracing data. We calculated the number of contacts per index case, the secondary attack rate (% contacts infected), and effective R number (new cases per index case), and used logistic regression to estimate how age, gender, and contact type affected transmission.

**Results:** Most contacts and transmission events were between family members. Contacts <40 years were less likely to be infected, while infected individuals >50 were more likely to die than younger patients. Contact tracing identified 917 index cases with mean 3.1 contacts tested per case, primarily family members. The overall secondary attack rate was 28% (95% confidence interval [CI]: 26-29%) and effective R number was 0.87 (95%CI 0.81-0.93), peaking at 1.1 (95%CI 0.98-1.2) during the period with strongest restrictions.

**Conclusion:** Georgia effectively controlled the COVID-19 epidemic in its early stages, although evidence does not suggest transmission was reduced during the strict lockdown period.

**Research in Context:** *Evidence before this study:* We searched PubMed and MedRxiv for papers reporting research using contact tracing data to evaluate the characteristics of the COVID-19 epidemic in any country. A number of analyses were identified from Asia, including China, Taiwan, Maldives, Thailand, South Korea, and India, but none from other regions other than one previous analysis conducted in Europe, focusing on the first two months of the COVID-19 epidemic in Cyprus. Studies evaluated number of contacts and different contact types, secondary attack rate, and effective R number. However, none of these studies compared characteristics between different time periods or under varied levels of non-pharmaceutical interventions or restrictions on social mixing.

*Added value of this study:* In this study, we use contact tracing data from Georgia from all cases identified in the first four months of the epidemic, as well as testing and hospitalization data, to evaluate the number and type of contacts, effective R number (new cases per index case), and secondary attack rate (proportion of contacts infected) in this population, and whether these measures changed before, during, and after the lockdown period. We also evaluated how the chance of transmission varied by type of index case and contact. Our results indicate that number of contacts remained relatively low throughout the study period, so although the secondary attack rate was relatively high (28%) compared to that seen in studies in Asia (10-15%), the effective R number was less than one overall, peaking at 1.1 (0.98-1.2) during the strictest lockdown period, with easing of restrictions corresponding to a lower effective R of 0.87 (0.77-0.97). Most transmission occurred between family members with transmission very low between co-workers, friends, neighbours, and medical personnel, indicating that the restrictions on social mixing were effective at keeping the epidemic under control during this period.

*Implications of all the available evidence:* Our study presents the first analysis of the successful control of a COVID-19 epidemic in a European country, indicating that despite a high secondary attack rate, reduction in contacts outside the home, and a well-timed lockdown, were able to keep transmission under control.

## Introduction

Georgia reported their first case of COVID-19 on 26th February 2020. As of 24^th^ June 2020, 917 confirmed cases and 14 deaths had been reported. Georgia ‘s limited spread of COVID-19 over this period could be attributable to early non-pharmaceutical interventions (NPI). In January 2020, Georgia established a national committee focused on COVID-19^1^. Interventions included closing schools (early March), closing borders and quarantining international arrivals (late 2020), lockdown of high affected areas (late March) and full national lockdown (30th March, including restrictions on events, closure of non-essential businesses and curfew), with restrictions gradually lifting from 27^th^ April^2,3^. Additionally, they implemented extensive contact tracing and testing, with all confirmed cases being hospitalised. There was also screening of essential workers. A national protocol on COVID-19 treatment and care was developed. Infectious disease and intensive care physicians in all COVID-19 clinics were trained in COVID-19 clinical management.

By end of June, over 100,000 real-time polymerase chain reaction (RT-PCR) tests for SARS-CoV-2 had been conducted (>30,000 tests per million population), with 125 tests undertaken per case identified (rate=0.8%); a positivity rate similar to the Netherlands and Switzerland at that time^4,5^. World Health Organization (WHO) criteria suggest a positivity rate <5% indicates that an epidemic is under control^6^. As of 24th June 2020, Georgia had 229 cases per million population, the lowest rate in Europe (Figure 1). They also had a lower COVID-19 related death rate (3.5 deaths per million population) than all European countries except the Faroe Islands, Gibraltar, and the Vatican^5^.

**Figure 1.**
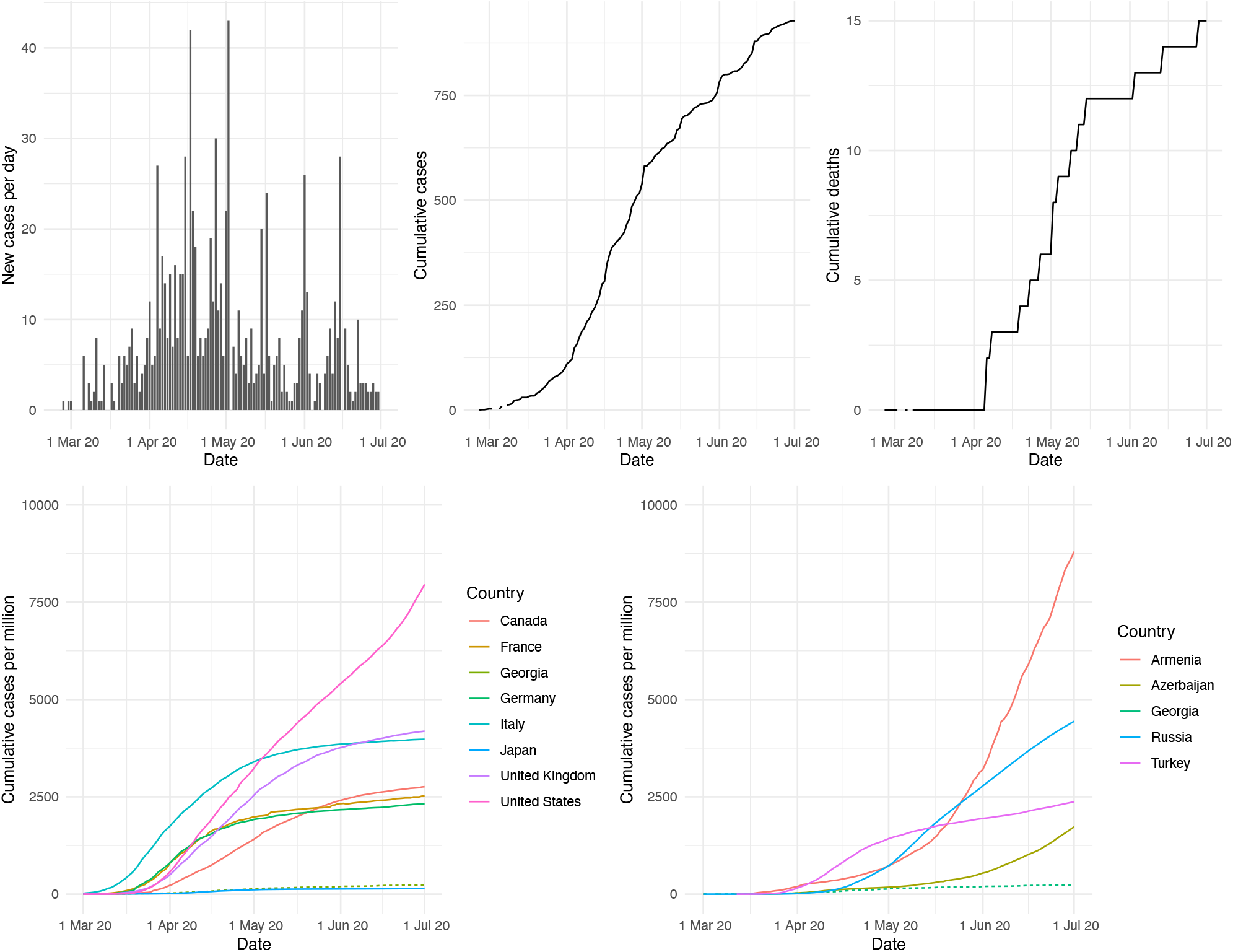
Trajectory of COVID-19 epidemic in Georgia and other countries. Top row, left: New cases per day in Georgia. Top row, middle: cumulative cases per day in Georgia. Top row, right: Cumulative deaths per day in Georgia. Bottom row, left: Cumulative cases per million population in Georgia (dashed line) and G7 countries. Bottom row, right: Cumulative cases per million population in Georgia (dashed line) and its bordering countries, data from Our World in Data ^5^.

Contact tracing, in which contacts of cases are identified, tested, and quarantined, is a core public health intervention used to control SARS-CoV-2 transmission ^7-10^. Analysing contact tracing data can also help understand the epidemiology of transmission in a particular setting, and determine the natural history of patients that might otherwise have not been tested, either because they are asymptomatic or did not access healthcare. Such analyses have already been conducted in several Asian settings including China^11-13^, Taiwan^14-16^, Maldives^17^, Thailand^18^, South Korea^19,20^, and India^21,22^, with only one previous analysis in Europe and Central Asia, focusing on the first two months of the COVID-19 epidemic in Cyprus ^23^.

Here we characterize the COVID-19 epidemic in Georgia through analysing data provided by the Georgian National Center for Disease Control (NCDC) on all PCR tests conducted (cases, routine testing and contract tracing). We compare the demographics of those testing positive and negative, and assess reported symptoms (fever or cough) and death rates by age. We also calculate the secondary attack rate for different types of contacts and number of secondary cases per primary case (effective reproductive number R), overall and by stage of lockdown in Georgia.

## Methods

### Data collection

COVID-19 testing started at NCDC ‘s Lugar Center reference laboratory from 4 February 2020. An electronic database was established for COVID-19 test results. Data were also collected on test and sample type, patient characteristics (gender, date of birth, geographic region/nationality), risk group, and symptoms (fever and cough only). Data was collected for all tested individuals, with further information collected for diagnosed patients, including length of hospitalization and additional symptoms. All confirmed cases were hospitalized during the study period.

Individuals were tested for COVID-19 for various reasons: symptoms according to the WHO standard definition, which were defined as: acute respiratory illness with fever and at least one other sign/symptom of respiratory disease, a history of travel to an area with local transmission or contact with a confirmed case; or severe acute respiratory infection requiring hospitalization and no other etiology that explains the clinical presentation^24^.

Non-symptomatic testing was conducted for contacts of confirmed cases; hospital patients or medical personnel with symptoms of respiratory disease; essential workers such as medical personnel, laboratory staff and transport/customs workers; beneficiaries and staff of care facilities for elderly and disabled; patients with early-stage tuberculosis (TB); and those in quarantine or self-isolation. Patients who met criteria for COVID-19 according to the WHO definition, but tested negative by RT-PCR with a specimen from an upper respiratory tract were defined as suspected cases, while waiting for recommended follow-up testing on a specimen from the lower respiratory tract.

In hospitalization data, cases were defined as either (i) mild - an acute respiratory infection without pneumonia; (ii) moderate - an acute respiratory infection with pneumonia but without shortness of breath; (iii) severe - Severe Acute Respiratory Infection (SARI) with pneumonia, difficulty breathing, sub-optimal oxygen saturation in blood; or (iv) critical - needing non-invasive or invasive ventilation, with other characteristics. Patients were discharged from hospital after normalization of temperature, improved radiographic examination of the chest, and virus clearance, with recovery defined as two consecutive negative PCR tests on upper/lower respiratory tract samples at least 24 hours apart.

For contact tracing, all confirmed case were contacted by phone to undertake a detailed epidemiological survey. All contacts were identified and contact information obtained. A contact is defined as any person who has had contact with a COVID-19 case from 48 hours before symptom onset to 12 days after. For an asymptomatic case, a contact is defined as someone who has had contact with the case from 48 hours before the sample which led to diagnosis was taken, to 12 days after. All contacts were called to inform them that they must isolate for 14 days, either at home or in a hotel.

Data on COVID-19 testing, contact tracing, symptoms, hospitalization, and outcomes were extracted and de-identified by NCDC, with a cutoff date of 24 June 2020. Patients were linked between testing, treatment and contact tracing databases using an anonymised unique identification number (ID).

### Analyses

We firstly evaluated whether the demographics of tested patients differed by test result (positive, negative, or suspected). This used the first test result recorded for each patient, excluding tests with no patient ID available. We also evaluated hospitalization data on disease severity and outcomes to consider differences by risk group, age group (0-49 compared to 50 or older) and gender. Differences were evaluated using the Kruskal-Wallis test (numeric variables) or Chi-squared test (categorical variables).

Secondly, we calculated the number of reported contacts per index case, the secondary attack rate (proportion of contacts infected) and effective R number (number of new infections per index case). Binomial 95% confidence intervals were calculated for the secondary attack rate, while Poisson confidence intervals were calculated for the effective R number assuming one unit of time between each case and contact. In the base-case we include all index cases in the calculation, including those identified through contact tracing, thus producing a measure of R over multiple generations of transmission. As a sensitivity analysis, we re-calculated R using only index cases who were not contacts, excluding individuals who could not be accurately matched due to missing IDs. In addition, as some contact test results were unknown, we present the secondary attack rate including the unknown outcomes in the denominator in the base-case, but excluded them in the sensitivity analysis.

Contact tracing data did not include dates for testing or symptom onset so we assumed that contacts were infected after their respective index cases. To calculate chains of transmission, we linked patients reported as both contacts and index cases, excluding those who could not be matched. Bivariate and multivariate logistic regression was used to evaluate whether age group (10-year groupings) and gender of index case or contact, and contact type were associated with contacts testing positive during the study period.

We lastly compared contact network characteristics, secondary attack rate, and effective R number for different time periods. We identified the first positive test date for each index case, then assigned each contact tracing network to one of three time periods according to the test date of the index case, either 26 February-29 March (initial restriction period), 30 March-26 April (full lockdown), or 27 April-24 June (lifting of restrictions). We used logistic regression to determine whether the odds of transmission differed in the three time periods. This built on the results of the previous logistic regression model, by including co-variates which had 95% confidence intervals not overlapping 1, and simplifying categorical variables where no difference was observed between some of the original categories.

Data analyses were conducted in R version 3.6.1 and no imputation was conducted for missing data so analyses were conducted on complete cases.

### Ethics

Ethical approval (exemption) was obtained for this study from the National Center for Disease Control (NCDC) institutional review board (IRB), under reference # 2020-027, 02 June 2020. The study was deemed exempt from IRB due to using retrospective anonymized data. The study was also registered with University of Bristol ‘s research governance database.

## Results

### Demographics of those tested (tested database)

Results were available for 96,213 RT-PCR tests, of which 95,504 (98.8%) were conducted on nasal swab samples, with the remainder conducted on blood (19), aspirate (10), or fecal (2) samples, with 1,128 (1.1%) missing sample type information. Approximately 2.0% (1,931) of test results were positive and 0.3% (260) suspected. Personal ID numbers were missing for 1,719 results, with the remaining 94,494 records contained 67,860 unique individuals, indicating that 28% of tests were repeat tests.

The breakdown of results for the first test for each unique patient is in Table 1. Positive results were marginally more likely in older patients but did not differ by gender. Patients testing positive were more likely to report fever than those testing negative, and symptomatic individuals were more likely to test positive than other risk groups.

**Table 1:** Age, gender, reported fever or cough, and risk group (reason for testing) for 67,860 uniquely identified individuals tested for SARS-CoV-2 by RT-PCR (first test result).

| Category | Test result |  |  |  | p value* |
| --- | --- | --- | --- | --- | --- |
|  | Negative<br>(N=66,941) | Positive<br>(N=763) | Suspected<br>(N=156) | Total (N=67,860) |  |
| **Age** |  |  |  |  | 0.065 |
| Mean (SD) | 41.3 (18.9) | 42.7 (19.8) | 42.7 (20.1) | 41.3 (18.9) |  |
| Missing | 9 | 0 | 0 | 9 |  |
| **Gender** |  |  |  |  | 0.547 |
| Female | 34,552 (51.6%) | 389 (51.0%) | 74 (47.4%) | 35,015 (51.6%) |  |
| Male | 32,389 (48.4%) | 374 (49.0%) | 82 (52.6%) | 32,845 (48.4%) |  |
| **Risk Group** |  |  |  |  | < 0.001 |
| Symptomatic | 20,045 (36.6%) | 634 (84.8%) | 63 (44.7%) | 20,742 (37.3%) |  |
| Healthcare worker | 11,719 (21.4%) | 7 (0.9%) | 13 (9.2%) | 11,739 (21.1%) |  |
| Quarantine or isolation | 11,540 (21.1%) | 43 (5.7%) | 42 (29.8%) | 11,625 (20.9%) |  |
| Contact of case | 4,057 (7.4%) | 50 (6.7%) | 13 (9.2%) | 4,120 (7.4%) |  |
| Transport/other essential worker | 3,064 (5.6%) | 8 (1.1%) | 7 (5.0%) | 3,079 (5.5%) |  |
| Non-COVID-19 patient | 1,460 (2.7%) | 1 (0.1%) | 2 (1.4%) | 1,463 (2.6%) |  |
| Other | 2,904 (5.3%) | 5 (0.7%) | 1 (0.7%) | 2,910 (5.2%) |  |
| Missing* | 12,152 (18.2% of N) | 15 (2.0% of N) | 16 (10.3% of N) | 12,193 (18.0% of N) |  |
| **Fever** |  |  |  |  | 0.006 |
| No | 6,003 (32.6%) | 30 (21.0%) | 7 (21.9%) | 6,040 (32.5%) |  |
| Yes | 12,405 (67.4%) | 113 (79.0%) | 25 (78.1%) | 12,543 (67.5%) |  |
| Missing* | 48,533 (72.5% of N) | 620 (81.3% of N) | 124 (79.5% of N) | 49,277 (72.6%) |  |
| **Cough** |  |  |  |  | 0.827 |
| No | 12,227 (68.9%) | 88 (66.7%) | 21 (67.7%) | 12,336 (68.9%) |  |
| Yes | 5,527 (31.1%) | 44 (33.3%) | 10 (32.3%) | 5,581 (31.1%) |  |
| Missing* | 49,187 (73.5%) | 631 (82.7%) | 125 (80.1%) | 49,943 (73.6%) |  |

P-value assesses probability of difference by test result evaluated by Kruskal-Wallis test (numeric variables) or Chi-squared test (categorical variables) excluding missing values.
\*For categorical variables, the % in each reported group are presented to add up to 100% to align with the statistical analyses, with the % missing presented separately. SD: Standard deviation

### Demographics of those infected (treated database)

Hospitalization data, comorbidities, and outcomes were available for 500 diagnosed patients hospitalized between February 25 and 2 May; no further data were available at the time of data extraction.

Women in this database were on average 6 years older than men (46 versus 40). Most (72%) were infected in Georgia. Disease severity and mortality varied by age, with 94.8% of patients (289/305) under 50 years having mild or moderate disease and one death (0.3%), compared to 76.4% of patients (149/195) 50 years or older with 12 deaths (6.2%) (Table 2). There was difference in severity or mortality by gender. The median hospital stay was 21 days, ranging 2-65 days.

**Table 2:** Demographics and disease severity of 500 diagnosed/hospitalized patients.

| Category | Age group |  |  | p value* |
| --- | --- | --- | --- | --- |
|  | 0-49 (N=305) | 50+ (N=195) | Total (N=500) |  |
| **Age (years)** |  |  |  | < 0.001 |
| Mean (SD) | 30.4 (12.6) | 62.1 (9.9) | 42.7 (19.3) |  |
| **Gender** |  |  |  | < 0.001 |
| Female | 132 (43.3%) | 121 (62.1%) | 253 (50.6%) |  |
| Male | 173 (56.7%) | 74 (37.9%) | 247 (49.4%) |  |
| **Disease severity** |  |  |  | < 0.001 |
| Mild | 208 (68.2%) | 67 (34.4%) | 275 (55.0%) |  |
| Moderate | 81 (26.6%) | 82 (42.1%) | 163 (32.6%) |  |
| Severe | 11 (3.6%) | 28 (14.4%) | 39 (7.8%) |  |
| Critical | 5 (1.6%) | 18 (9.2%) | 23 (4.6%) |  |
| **Length of hospital stay** |  |  |  | 0.616 |
| Median (IQR) | 20 (16, 27) | 21 (17, 27) | 21 (16, 27) |  |
| Missing | 1 | 0 | 1 |  |
| **Outcome** |  |  |  | < 0.001 |
| Died | 1 (0.3%) | 12 (6.2%) | 13 (2.6%) |  |
| Recovered | 304 (99.7%) | 183 (93.8%) | 487 (97.4%) |  |
| **Place of exposure** |  |  |  | 0.024 |
| Georgia | 213 (69.8%) | 149 (76.4%) | 362 (72.4%) |  |
| Europe and Central Asia excluding Georgia | 42 (13.8%) | 12 (6.2%) | 54 (10.8%) |  |
| North America | 4 (1.3%) | 3 (1.5%) | 7 (1.4%) |  |
| Middle East and North Africa | 4 (1.3%) | 0 (0.0%) | 4 (0.8%) |  |
| Latin America and the Caribbean | 0 (0.0%) | 1 (0.5%) | 1 (0.2%) |  |
| Missing | 42 (13.8%) | 30 (15.4%) | 72 (14.4%) |  |

\*P-value assesses probability of difference by age group (0-49 or 50+ years) evaluated by Kruskal-Wallis test (numeric variables) or Chi-squared test (categorical variables) excluding missing values. SD: Standard deviation; IQR: interquartile range

### Contact network

The contact tracing database contained 2,882 links between 917 index cases and contacts that were tested for COVID-19. Each index case had on average 3.1 contacts that were tested (median 2, range of 1-30; Table 3). Family members made up most of the reported contact types in each time period, with contacts and transmission varying by age within families (Supplementary Figure 1). Of all 2,882 contacts reported, 312 were also included as index cases, due to missing patient IDs for both cases (15% unique IDs missing) and contacts (28% unique IDs missing). When contacts without patient IDs are excluded, there are 252 unconnected sub-networks within the contact tracing data, with the longest transmission chain having 4 links.

**Table 3:** Summary of index cases and contacts reported over three time periods: initial restriction period (26 February to 29 March), full lockdown (30 March to 26 April), and gradual easing of restrictions (from 27 April to 24 June), and overall (for comparison, not included in statistical comparison).

|  | Initial restriction period | Lockdown | Restrictions easing | Overall* | p-value |
| --- | --- | --- | --- | --- | --- |
| Index cases | 66 | 352 | 342 | 917 |  |
| Total contacts | 180 | 1296 | 1048 | 2882 |  |
| **Network measures** |  |  |  |  |  |
| Median contacts per case | 2 | 3 | 2 | 2 | <0.001<sup>2</sup> |
| Mean contacts per case | 2.7 | 3.7 | 3.1 | 3.1 |  |
| Range of contacts per case | 1-17 | 1-23 | 1-30 | 1-30 |  |
| **Total contacts by type** |  |  |  |  | <0.001<sup>1</sup> |
| Family | 102 (56.7%) | 803 (62.0%) | 682 (65.1%) | 1766 (61.3%) |  |
| Co-worker | 11 (6.1%) | 201 (15.5%) | 136 (13.0%) | 397 (13.8%) |  |
| Friend | 28 (15.6%) | 57 (4.4%) | 42 (4.0%) | 148 (5.1%) |  |
| Neighbor | 15 (8.3%) | 142 (11.0%) | 34 (3.2%) | 212 (7.4%) |  |
| Medical personnel | 8 (4.4%) | 19 (1.5%) | 36 (3.4%) | 63 (2.2%) |  |
| Unknown | 16 (8.9%) | 74 (5.7%) | 118 (11.3%) | 296 (10.3%) |  |
| **Transmission** |  |  |  |  |  |
| Secondary attack rate | 21.1% (15.4-27.8%) | 29.6% (27.1-32.1%) | 28.3% (25.6-31.2%) | 27.6% (26.0-29.3%) |  |
| Effective R number | 0.58 (0.41-0.79) | 1.1 (0.98-1.2) | 0.87 (0.77-0.97) | 0.87 (0.81-0.93) |  |
| Cases in contacts | 38 | 383 | 297 | 795 |  |

<sup>1</sup>Pearson's Chi-squared test for difference between three time periods. <sup>2</sup>Kruskal-Wallis test for difference between three time periods. \*Overall estimate not included in statistical comparison, and individuals included in overall who were not matched to one of the three time periods.

Overall, 795 contacts tested positive, with no test outcomes available for 296 (10%) of contacts. If individuals with missing outcome data are included in the denominator (i.e. assumed not to be a case), the overall secondary attack rate was 28% (795/2882, 95% confidence interval (CI) 26-29%) and effective R number was 0.87 (795/917, 95%CI 0.81-0.93). However, if they are excluded from the calculation, the secondary attack rate increases to 31% (795/2586, 95%CI 29-33%). With only the 477 index cases not included as contacts, there were 1,342 reported contacts with 301 diagnosed as secondary cases. This gives a secondary attack rate of 22% (301/1342, 95%CI 20-25%) and R number of 0.63 (301/477, 95%CI 0.56-0.71).

### Variables associated with transmission

In bivariate analysis, the strongest association for a contact testing positive was with the type of contact, with transmission between family members (reference group) being more likely than between co-workers (OR 0.52, 95%CI 0.40-0.66), friends (OR 0.40, 95%CI 0.25-0.59), neighbours (OR 0.17 95%CI 0.10-0.26), or medical personnel (no cases, N=63). Results changed little in adjusted analyses (Figure 2).

**Figure 2.**
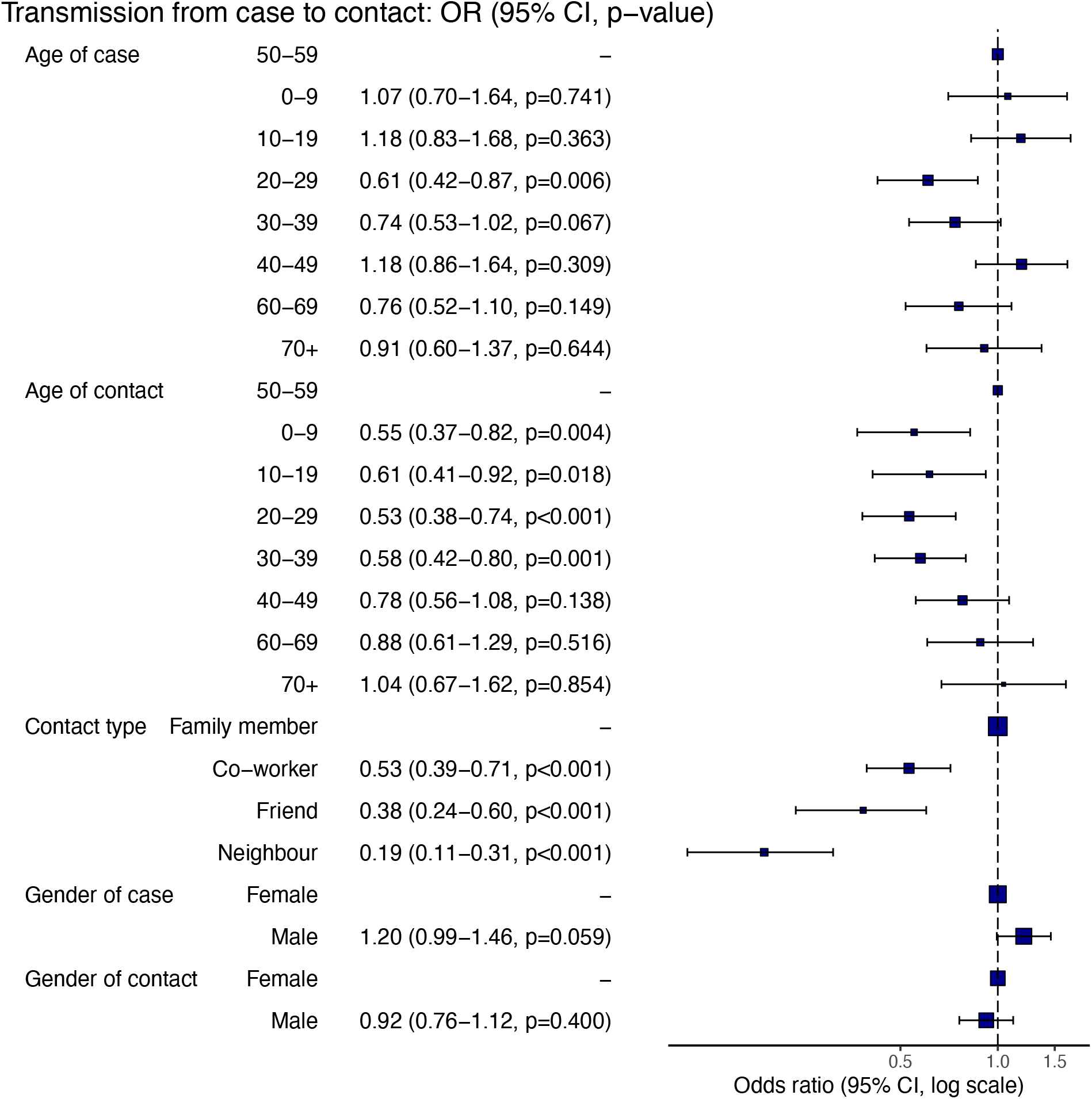
Adjusted odds ratios (ORs) and 95% confidence intervals (CI) for COVID-19 diagnosis among close contacts. Group-specific attack rates, risk factors, and unadjusted ORs are shown in Supplementary
Table 1.

Contacts under 50 years have lower odds of testing positive than those 50 or older. The results are similar in the adjusted analysis (Figure 2), with contacts younger than 40 having lower odds of testing positive than those in their 50s. For the index cases, only those aged 20-29 have lower odds of infecting their contacts (aOR 0.61, 95%CI 0.42-0.87) than those aged 50-59.

The gender of the index case was strongly associated with a contact testing positive in the bivariate analysis, with contacts of males being more likely to test positive than contacts of females (OR 1.25, 95%CI 1.06-1.48), however, this association weakened after adjustment (aOR 1.21, 95%CI 0.99-1.46). The gender of the contact was not associated with testing positive.

### Transmission by time period

We were able to identify the first positive testing date for 760 index cases. In the initial restriction period, there were 66 cases, with 352 in the full lockdown period, and 342 during the lifting of restrictions phase (Table 3). The R number varied by time period, being highest during lockdown (1.1, 95%CI 0.92-1.2) and lowest in the initial restriction period (0.58, 95%CI 0.41-0.79). The secondary attack rate varied less by time period.

In bivariate analysis, the odds of transmission from a case to a contact were lowest in the initial restriction period (OR 0.66, 95%CI 0.45-0.96, reference lockdown), with no difference between the lockdown and restrictions easing periods (OR 1.03 95%CI 0.86-1.23) (Supplementary Table 2). However, when contact type (family or other) and age of contact (up to 40 years old vs 40 years and older grouped based on initial analysis) were adjusted for in the analysis, the odds of transmission is the same in the initial restriction period compared to lockdown (aOR 0.85, 95% CI 0.55-1.31), with slighty lower odds of transmission when restrictions were eased compared to lockdown (aOR 0.79, 95% CI 0.64-0.97) (Figure 3).

**Figure 3.**
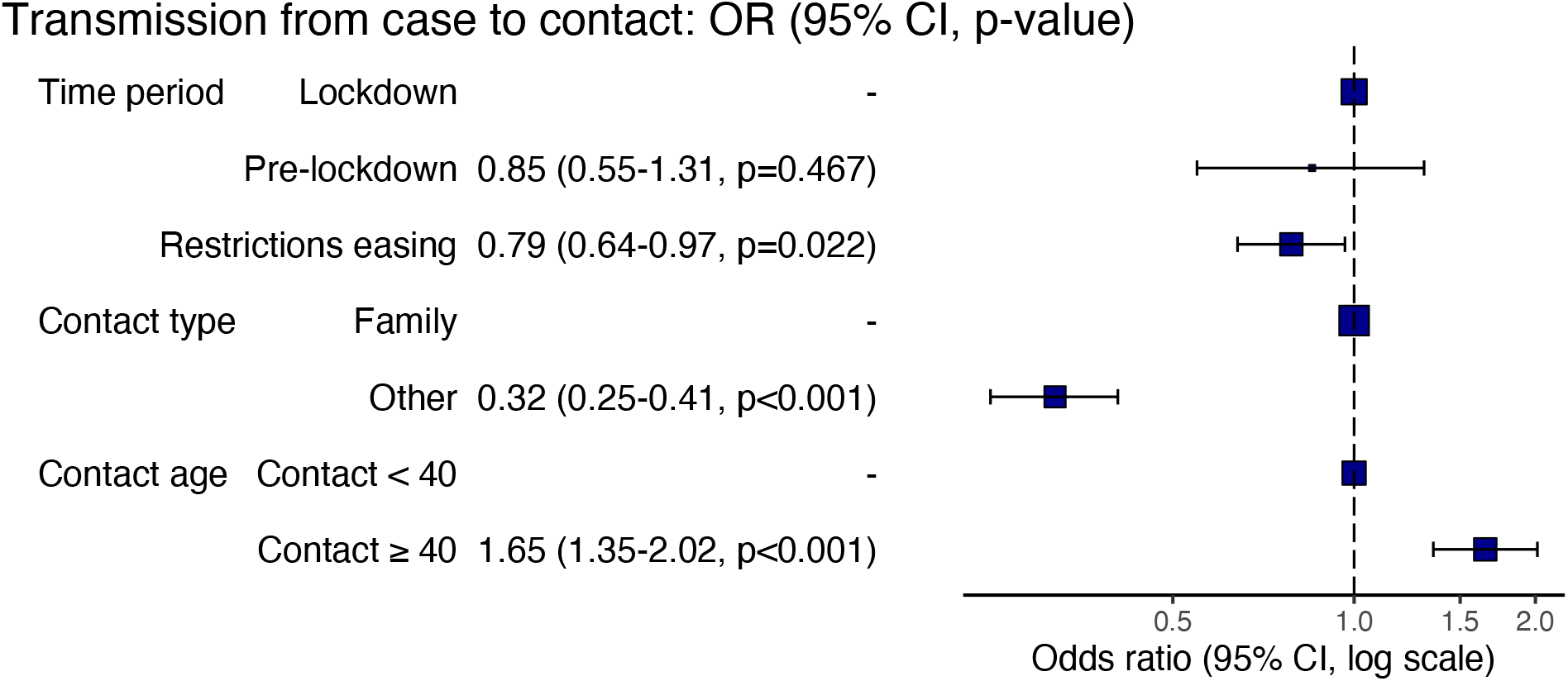
Adjusted odds ratios (ORs) and 95% confidence intervals (CI) for COVID-19 diagnosis among close contacts by time period. Group-specific attack rates, risk factors, and unadjusted ORs are shown in Supplementary Table 2.

Behaviour varied by time period (Table 3), with the number of contacts reported by each index case being slightly higher in lockdown, perhaps reflecting a difference in reporting or the type of people who were infected during this period. This is supported by differences in types of contacts observed during each period, with the proportion of contacts being co-workers increasing, while proportion being friends decreasing, from the initial restriction to lockdown periods.

## Discussion

This study evaluated data on cases and their contacts in the early stage of the COVID-19 epidemic in Georgia. Over this time, case numbers remained low, in contrast to other countries, including those bordering Georgia. In this study, we found diagnosed cases were equally likely to be men or women and had a similar age to those that tested negative. However, of those infected and hospitalized, more were men, women were older than men (by 6 years), and patients over 50 years old were more likely to die than younger patients (6.2% older patients and 0.3% of younger patients died). Overall, the effective R number of 0.87 confirms that at this point the epidemic was under control and not growing exponentially. The majority of contacts and transmission events were between family members, which also had a higher odds of transmission. Conversely, contacts younger than 40 had 65% lower odds of being infected, while male cases and those between 20-29 were less likely to infect their contacts. Transmission didn ‘t decrease during the full lockdown period, possibly due to changes in the type of contacts resulting from the implemented restrictions.

The majority of positive cases were identified through symptomatic testing (>80%), with most others found in quarantining patients or contacts of positive cases. Unfortunately, details of the specific symptoms experienced were missing/not reported for many individuals with ‘symptoms ‘, making it difficult to draw firm conclusions about which symptoms were experienced by positive cases. Otherwise, very few positive cases were identified through routine screening of healthcare workers, while no transmission was recorded between healthcare personnel and patients. This suggests a low risk of COVID-19 transmission to healthcare workers, in agreement with a systematic review of secondary attack rate in healthcare settings which estimated it to be 0.7%^25^.

The high number of family contacts in our study likely reflects the restrictions put in place to achieve social distancing, with non-essential businesses and schools closing, and most people working from home. However, the proportion of contacts that were family members only increased slightly during lockdown, while the proportion that were co-workers actually increased. Indeed, unexpectedly, the average number of contacts was also highest during lockdown. Although this could be due to people being more aware of their social contacts during the lockdown period, it could also reflect the heightened risk experienced by essential workers as well as family groups being in closer contact. The effective R number and secondary attack rate were also highest during the lockdown period, with R slightly above 1 indicating onwards transmission. R then decreased to 0.87 when restrictions were eased. Although this is counterintuitive, it may reflect that changes in policies were implemented at the correct time, such that lockdown kept R from rising, and that easing of restrictions occurred once R was decreasing. It is also important to note that the easing restrictions period still included many restrictions and was not a return to low level of restrictions seen before the lockdown.

### Comparison to other studies

Contact tracing is a key public health response to infectious disease, which evidence suggests was implemented effectively in Georgia. In many other settings, including the USA and UK, it has not been a main focus of their COVID-19 response ^26^. Modelling indicates that contact tracing requires considerable investment to be effective, with national-level coordination being required^27,28^. Contact tracing alone will not control a disease if people do not report their contacts and/or it is difficult to reach their contacts^29^. Georgia has a relatively small population (∼4 million people) with a strong national disease control agency, which likely increased the feasibility of undertaking effective contact tracing at a national level. In addition, contact tracing is most effective when case numbers are low because direct transmission events are more likely to be identified. Through acting quickly, Georgia ‘s overall strategy seemed to ensure this occurred.

In this study, we observed a case to contact transmission rate (secondary attack rate) of 28% which is much higher than observed in other contact tracing studies. The probability of infection of close contacts (eg household/physical contact) was found to be 10-15% in Cyprus, India, China, South Korea and Thailand^11,18,19,22,23^. In addition, a recent systematic review of household-specific secondary attack rates found an overall rate of 18%, though this varied by study from 34-55%^25^. None of the previous studies compared results over time or across different levels of restrictions.

The higher rate found in this study could indicate an underreporting of close contacts or negative test results, or may reflect differences in COVID-related contact restrictions and/or a heightened transmission risk within households due to social or cultural differences. Unfortunately, no other comparable studies exist from Eastern Europe or Central Asia, while other contact tracing studies only performed confirmatory testing on symptomatic contacts^14,30^. Interestingly, though, despite having a high secondary attack rate, the effective R was still low in Georgia due to low numbers of contacts, mean of 3.1, compared to 7.3 in India^22^ and 13.3 in China^11^, though strict lockdowns were not implemented in those settings.

### Strengths and limitations

This study used national level public health data to evaluate how COVID-19 spread in the early stages of the epidemic in Georgia. A large number of tests were conducted and reported, providing a rich resource. However, some variables, such as specific symptoms (fever or cough), were underreported in the testing data, even among those tested for being symptomatic, with higher rates of missing data for those testing negative. This could lead to collider bias that distorts the association between variables, for example if a risk factor and outcome both have an impact on whether data are recorded ^31^. Furthermore, missing patient IDs in the contact tracing data may have led us to underestimate the transmission chains because some positive contacts could not be linked to others. In addition, only one reason for testing was given even though some patients might have fallen into multiple categories (e.g. an essential worker experiencing symptoms). Lastly, any hospitalization data was only available for the first 500 patients with disease outcomes, either discharge or death, and so we may have underrepresented more severe patients who were hospitalized but had not as yet one of these outcomes.

## Conclusions

The implementation of widespread contact tracing with non-pharmaceutical interventions to reduce contact between people while allowing essential work to continue seems to have been effective at controlling the early epidemic in Georgia. Indeed, their respone demonstrated the strength of their public health system in terms of diagnostic capability and supportive care for COVID-19 cases. This was likely helped by recent investments related to the national initiative to eliminate Hepatitis C virus, including improvements in diagnostic facilities and training pathways for primary care doctors^32^. Unfortunately, though, from September 2020 case numbers have increased drastically in Georgia, with only 1,510 cumulative cases reported up to 1 September but nearly 228,000 cases by end of the year^33^. This has led to a change in strategy, as existing contact tracing strategies could not keep up with the rapid increase in cases, and mild cases are no longer hospitalized. Future epidemiological and modelling research is needed to understand how the COVID-19 epidemic in Georgia was kept under control in the early period, but not in the subsequent period. This could lead to important insights for how to design future containment strategies in Georgia and elsewhere.

## Data Availability

Data used in the study will be made available upon publication, including the de-identified contact database, de-identified hospital outcome data, and de-identified test data.

## Supplementary Information

**Supplementary Table 1:**
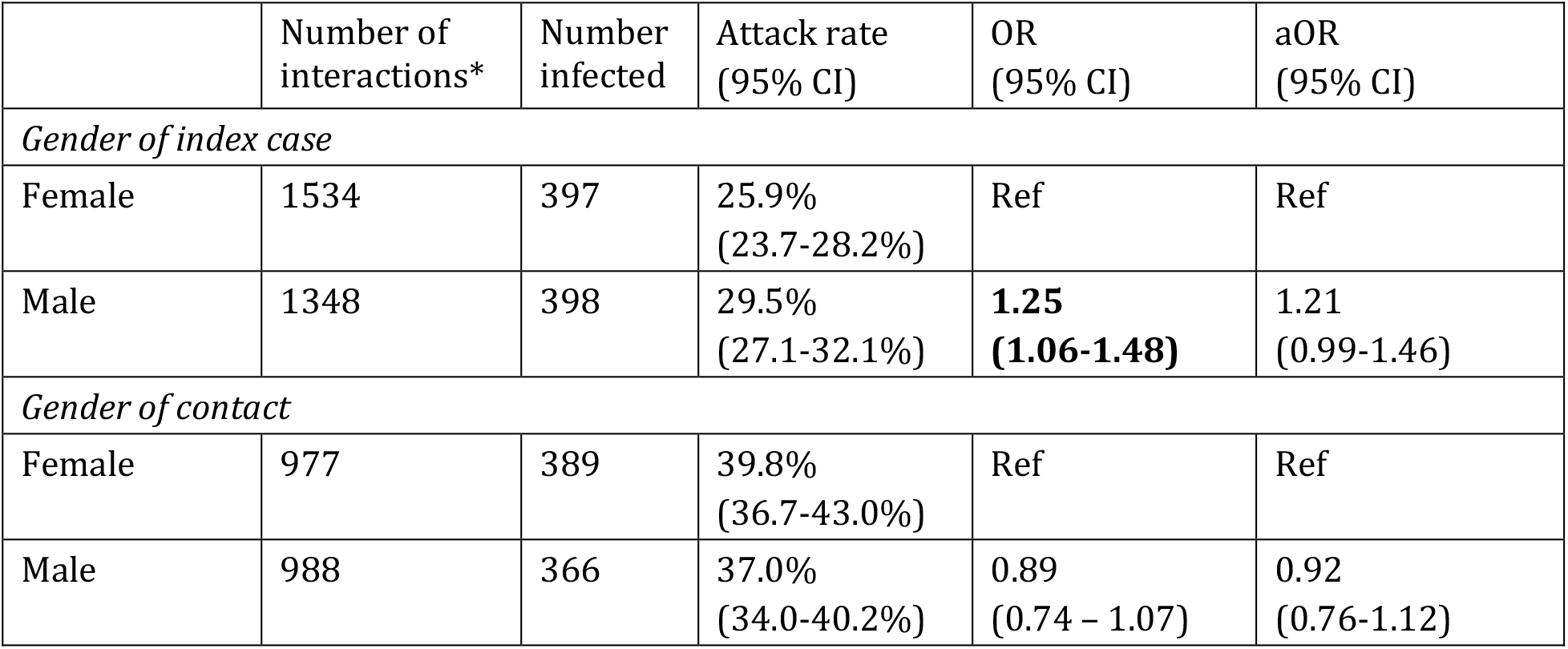
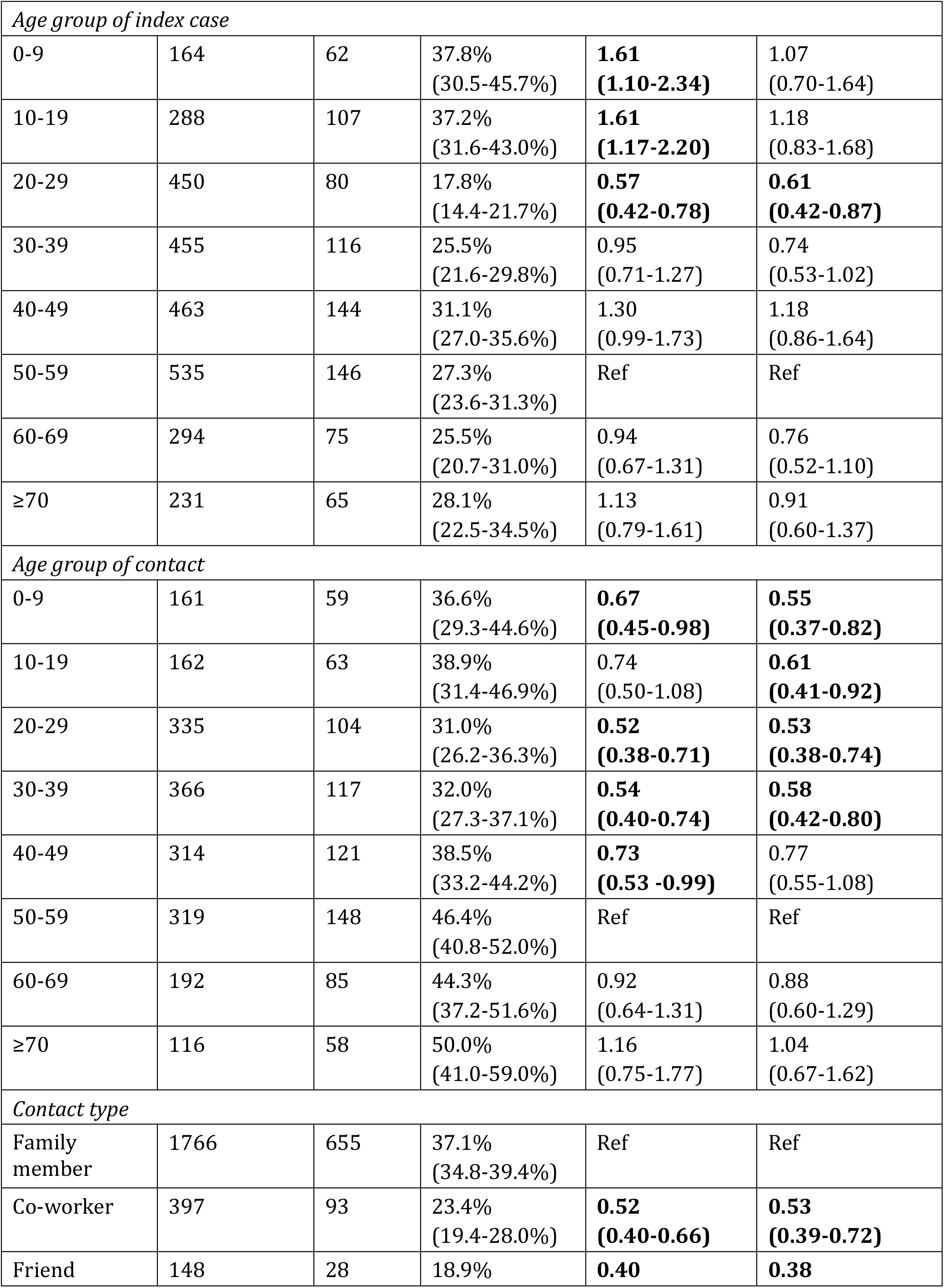
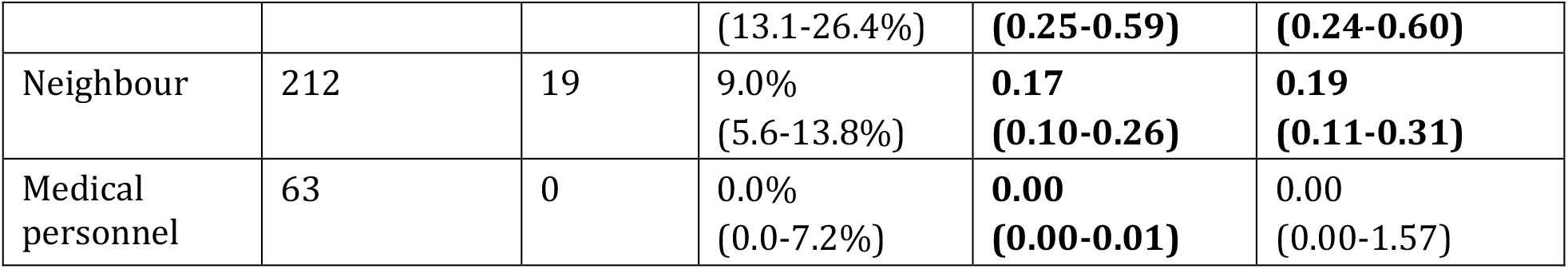
Group-specific attack rates, risk factors, adjusted and unadjusted ORs for COVID-19 diagnosis among close contacts. *number of interactions for which the variable is available. Odds ratio (OR) 95% confidence intervals (CI) not overlapping 1 are shown in bold font.

**Supplementary Table 2:** Group-specific attack rates, adjusted and unadjusted ORs for COVID-19 diagnosis among close contacts for risk factors identified in full analysis and time period (initial restriction period, lockdown, or restrictions easing) *number of interactions includes those with unknown test result. Odds ratio (OR) 95% confidence intervals (CI) not overlapping 1 are shown in bold font.

|  | Number of interactions* | Number infected | Attack rate (95% CI) | OR (95% CI) | aOR (95% CI) |
| --- | --- | --- | --- | --- | --- |
| *Contact age group* |  |  |  |  |  |
| <40 years old | 1024 | 343 | 33% (31-36%) | Ref | Ref |
| ≥40 years | 941 | 412 | 44% (41-47%) | **1.55 (1.29-1.86)** | **1.65 (1.35-2.02)** |
| *Contact type* |  |  |  |  |  |
| Family | 1766 | 655 | 37% (35-39%) | Ref | Ref |
| Other | 1116 | 140 | 13% (11-15%) | **0.35 (0.38-0.43)** | **0.32 (0.25-0.41)** |
| *Time period* |  |  |  |  |  |
| Initial restriction period | 180 | 38 | 21% (16-28%) | **0.66 (0.45-0.96)** | 0.85 (0.55-1.31) |
| Lockdown | 1296 | 383 | 28% (26-31%) | Ref | Ref |
| Restrictions easing | 1048 | 297 | 30% (27-32%) | 1.03 (0.86-1.23) | **0.79 (0.64-0.97)** |

**Supplementary Figure 1:**
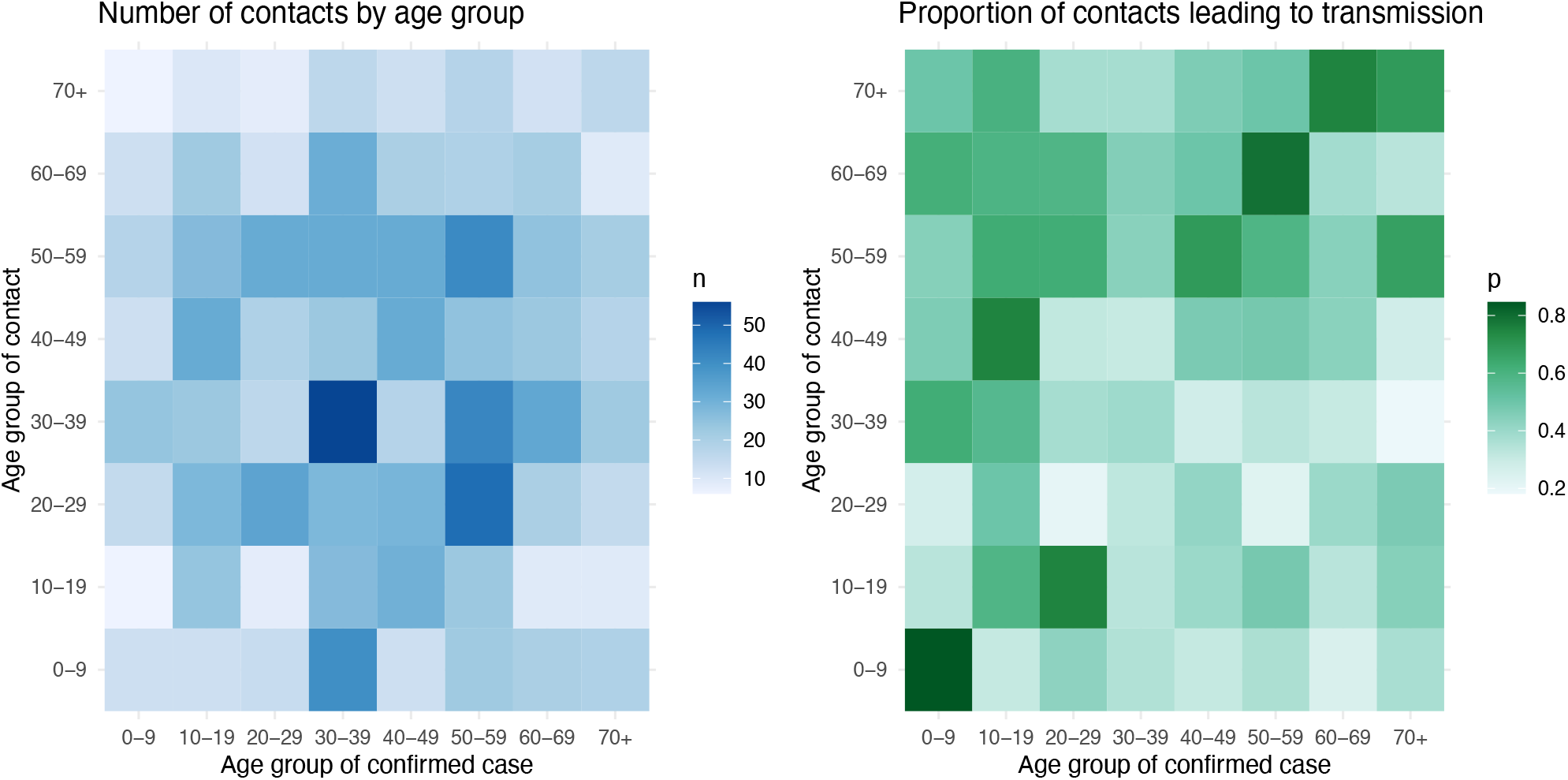
Heat map of within-family contacts between age groups (left) and proportion of within-family contacts leading to transmission by age group (right).

